# Geospatial Impact Indexing of Agricultural Incidents: A Multi-Criteria Risk Assessment in the U.S. Midwest

**DOI:** 10.64898/2026.05.06.26352581

**Authors:** Ege Duran, Omer Mermer, Ibrahim Demir

## Abstract

Traditional agricultural safety assessments often rely on raw incident counts that emphasize exposure but underrepresent outcome severity. This study presents a multi-criteria impact framework to distinguish frequency-driven activity patterns from severity-driven risk across the U.S. Midwest. Agricultural incident records from 2012 to 2023 across seven states were analyzed using descriptive statistics, county-level mapping, and quartic kernel density estimation. Comparative impact indices were constructed using Analytic Hierarchy Process (AHP) and Geometric-Fuzzy AHP weighting schemes to integrate incident frequency, outcome severity, and post-incident survivability. Results indicate that while overall incident frequency is strongly concentrated in northwestern Iowa, reflecting intensive agricultural activity, fatal outcomes exhibit a broader spatial footprint extending across central and northern Iowa and into central–southern Minnesota. Severity-weighted mapping further consolidates northwestern Iowa and the Minnesota–Iowa corridor as dominant high-impact zones. At the regional scale, Geometric-Fuzzy AHP produced consistently lower mean scores and reduced dispersion than AHP, yielding smoother spatial gradients while preserving the primary hotspot structure. These findings demonstrate that frequency-based mapping alone fails to capture the multi-dimensional nature of agricultural risk. By explicitly linking incident locations with survival infrastructure, this research provides an evidence-based framework for targeting safety interventions and improving rural emergency medical service planning.

## 1. Introduction

Agriculture consistently ranks as one of the most hazardous industries in the United States, presenting persistent risks to operators, workers, and family members (New-Aaron et al., 2019). According to the Bureau of Labor Statistics, agriculture often records the highest rate of fatal occupational injuries, reported as 23.1 fatalities per 100,000 full-time equivalent workers in recent years, and significantly high rates of non-fatal injuries (U.S. Bureau of Labor Statistics, 2024; NIOSH, 2022). Recent literature emphasizes that the burden of agricultural injury is far more expansive and complex than simple casualty counts suggest, encompassing profound economic losses, long-term disability, and strain on rural healthcare systems (Scott & Sorenson, 2024). This challenge is particularly pronounced in the Midwest region, a hub of intensive agricultural production where machinery operations, livestock handling, and seasonal workloads create a complex landscape of occupational hazards (Ouattara et al., 2023; Johnson et al., 2021).

The risk profile of the Midwest is distinct and multifaceted. Studies focused on Iowa and surrounding states have identified specific operational risk factors that drive injury rates, including working more than 50 hours per week, the presence of large livestock, and the use of regular medications by an aging workforce (Jadhav et al., 2016; Sprince et al., 2003). Furthermore, “human factors” such as fatigue, hurry, and stress have been cited as primary contributors to incidents, often precipitating injuries related to falls, animal handling, and machinery operation (Jadhav et al., 2016; Jadhav et al., 2015). Despite the identification of these risk factors, effective prevention is hindered by the fragmented nature of injury surveillance. As noted by Gilblom et al. (2024), relying on single data sources often leads to significant underreporting and a skewed understanding of safety trends. Comprehensive surveillance requires integrating multiple data streams to overcome these limitations and accurately capture the scope of incidents across jurisdictional boundaries.

In parallel with these improvements in monitoring and data collection, advanced data-driven methodologies leveraging Machine Learning (ML) and Deep Learning (DL) have emerged for prediction in environmental and public health studies (Bayar et al., 2009; Krajewski et al., 2021). Recent studies have successfully applied different ML algorithms, such as XGBoost and Random Forest, to model complex, non-linear interactions between accident variables, offering higher predictive accuracy than traditional statistical methods (Khairuddin et al., 2023; Mermer et al., 2025; Ahmed et al., 2023). Furthermore, the integration of SHapley Additive exPlanations (SHAP) has addressed the interpretability challenges of these black box models, allowing researchers to quantify the precise contribution of specific features, such as operator age, type of injury agent, or safety gear usage, to injury outcomes (Wei et al., 2023; Mermer et al., 2026). However, while these data-driven approaches provide robust predictive insights into incident mechanics, they predominantly focus on event-level characteristics and often function independently of the macro-level environmental context, specifically, the spatial availability of emergency resources.

The severity of agricultural incidents is often exacerbated by the unique lone worker nature of the industry. Many incidents occur in remote locations where victims are working alone, creating significant barriers to immediate discovery and assistance (Etienne et al., 2023). In these scenarios, the golden hour, the critical window for medical intervention, is frequently compromised by structural deficits in rural infrastructure. Research has shown that survival outcomes for trauma victims are strongly correlated with the time taken to access emergency medical services (EMS), yet rural agricultural areas often face extended response times due to distance and the reliance on volunteer-based first responders (Etienne, 2024a). Furthermore, the ability to call for help is increasingly dependent on cellular communication infrastructure. Recent single-state analyses have highlighted that communication deserts, areas with insufficient cellular tower proximity, can delay EMS activation, potentially turning survivable injuries into fatalities (Etienne et al., 2024b).

Despite the recognized importance of spatial and infrastructural factors, many existing agricultural safety assessments continue to rely on survey-based data or isolated single-state analyses. For example, recent studies have characterized injury risk factors and body-part severity among Midwest farmers using surveillance surveys, yet these approaches often lack the geospatial resolution necessary to capture the environmental context of individual incidents (Ouattara et al., 2023). Similarly, while proximity-based analyses linking injuries to emergency medical services (EMS) and cellular infrastructure have been conducted at the single-state level, such as in Indiana, comprehensive multi-state frameworks remain limited (Etienne et al., 2024b).

More broadly, geospatial risk assessment has become a widely adopted approach for evaluating how hazard exposure and accessibility constraints interact across space (Alabbad et al., 2024; Duran et al., 2025a). Prior regional studies in the Midwest have demonstrated the utility of GIS-based approaches for evaluating community-level mitigation strategies, critical facility exposure, and infrastructure suitability under environmental stressors (Duran & Demir, 2026). This progress is progressed by the development of open-source programming frameworks for specialized environmental analysis and the use of interactive 3D serious games to enhance disaster awareness and mitigation education (Ramirez et al., 2022; Demiray et al., 2023). However, comparable multi-state geospatial applications remain limited in agricultural safety research, where incident consequences depend not only on exposure but also on spatial access to emergency and communication resources.

To address this gap, this study presents a geospatial analysis of agricultural incidents in the U.S. Midwest Region, specifically covering seven key states: Iowa, South Dakota, North Dakota, Nebraska, Missouri, Minnesota, and Kansas. Unlike previous studies that may focus solely on injury mechanics or localized infrastructure, this research builds a unified multi-state incident database spanning from 2012 to 2023. This study aims to identify and visualize the contextual relationships between incident locations and critical survival factors, including land use/land cover (LULC), farm-type characteristics, cellular communication infrastructure, and access to emergency services.

Furthermore, this study introduces a novel methodological framework for risk assessment by developing a multi-criteria incident impact index. AHP is a structured decision-making technique that decomposes complex problems into a hierarchy, using pairwise comparisons to derive priority weights for competing criteria (Saaty, 1987). However, traditional AHP can sometimes fail to capture the vagueness and uncertainty inherent in real-world human judgments. Fuzzy AHP (FAHP) addresses this by employing fuzzy set theory, allowing decision-makers to use linguistic variables (e.g., strongly important) rather than crisp numbers, thus providing a more realistic modeling of ambiguity and risk (Chang, 1996). These methods are essential for this study because agricultural risk is multidimensional; a high-impact incident is not defined solely by injury severity, but by the convergence of hazardous machinery, environmental isolation, and temporal constraints.

While AHP and FAHP have been widely applied in industrial safety and construction risk management, their application in U.S. agricultural safety remains very limited. For instance, researchers have successfully used AHP to prioritize safety climate factors in construction (Lim et al., 2021) and to assess machinery safety risks in manufacturing by integrating environmental and social criteria (Pačaiová et al., 2024). Within the Midwest specifically, these multi-criteria methods have recently gained traction for assessing environmental and infrastructural vulnerabilities. Integrated AHP and Fuzzy AHP frameworks have been used to evaluate the susceptibility of Iowa’s extensive bridge network to flood-induced damage and to develop state-level multidimensional agricultural drought risk maps (Duran et al., 2025b; Islam et al., 2025).

Further studies have utilized FAHP to map flood susceptibility in urban centers like Cedar Rapids and across various Iowa sub-basins by combining geophysical and socioeconomic factors (Cikmaz et al., 2023). Additionally, geospatial assessments of essential facilities, including hospitals, fire stations, and schools, have highlighted the critical link between infrastructure functionality, restoration time, and regional disaster resilience (Grant et al., 2024). In the agricultural sector, recent international studies have employed Fuzzy AHP to identify critical risk factors in project investments and to rank biomechanical hazards for farm workers, demonstrating the method’s capacity to handle the complex, data-scarce nature of agricultural environments (Zandi et al., 2020; Khadem & AlAli, 2024). By adapting these frameworks to the Midwest context, this study provides a novel, quantitative mechanism to assess the total impact of agricultural incidents beyond simple fatality counts.

By utilizing the AHP and FAHP, this research moves beyond simple frequency counts to quantify the impact of incidents based on a weighted integration of severity, hazard intrinsic to the type of incident (e.g., tractor rollovers, grain entrapment), temporal relevance, and spatial accessibility to care. To our knowledge, no previously conducted study has performed such a comprehensive geospatial analysis and multi-criteria risk assessment covering these seven states. By connecting incident locations with the survival infrastructure of the Midwest, this study provides a robust evidence base for targeting safety interventions, improving rural EMS planning, and addressing the digital divide in agricultural safety.

The contributions of this study are threefold. First, it constructs a unified, multi-state geospatial incident database using 2012–2023 surveillance data from Central States Center for Agricultural Safety and Health (CS-CASH), providing a comprehensive safety profile for the seven-state Midwest region while overcoming the limitations of fragmented, single-state reporting. Second, it delivers a granular geospatial analysis of survival infrastructure, identifying critical safety gaps by explicitly overlaying incident locations with cellular communication coverage and emergency medical service (EMS) accessibility. This spatial integration allows for the quantification of vulnerability specifically for lone workers in remote agricultural environments. Third, the study develops and applies a novel multi-criteria incident impact index utilizing both the Analytic Hierarchy Process (AHP) and Geometric-FAHP. This methodological framework moves beyond traditional frequency-based metrics to provide a robust, multidimensional assessment of agricultural incidents, enabling a clearer distinction between exposure-driven activity patterns and severity-driven risk concentrations across the region.

The remainder of this paper is organized as follows: Section 2 describes the research methodology, including geospatial data preprocessing and the development of the multi-criteria impact framework. Section 3 presents the results and discussion, including data analysis, geospatial clustering patterns, and comparative impact density mapping. Section 4 concludes with major findings, policy implications, and directions for future research.

## 2. Methodology

This study builds a unified multi-state geospatial incident database (2012–2023), produces a descriptive spatiotemporal profile, and then develops a multi-criteria incident impact index using AHP and Geometric-FAHP.

### 2.1. Study Area

The study area comprises seven states in the U.S. Midwest (Iowa, Kansas, Minnesota, Missouri, Nebraska, North Dakota, and South Dakota) representing a region of global significance for food and fuel production (Gelfand et al., 2013; Wright & Wimberly, 2013). This region is characterized by intensive row-crop production, extensive livestock operations, and a diverse range of agricultural land use/land cover (LULC) profiles that contribute to its unique occupational risk landscape (Ouattara et al., 2023; Sprince et al., 2003). As shown in Figure 1, the agricultural composition of this region has remained relatively stable in its core but has experienced notable shifts in peripheral crop dominance over the last decade.

**Figure 1:**
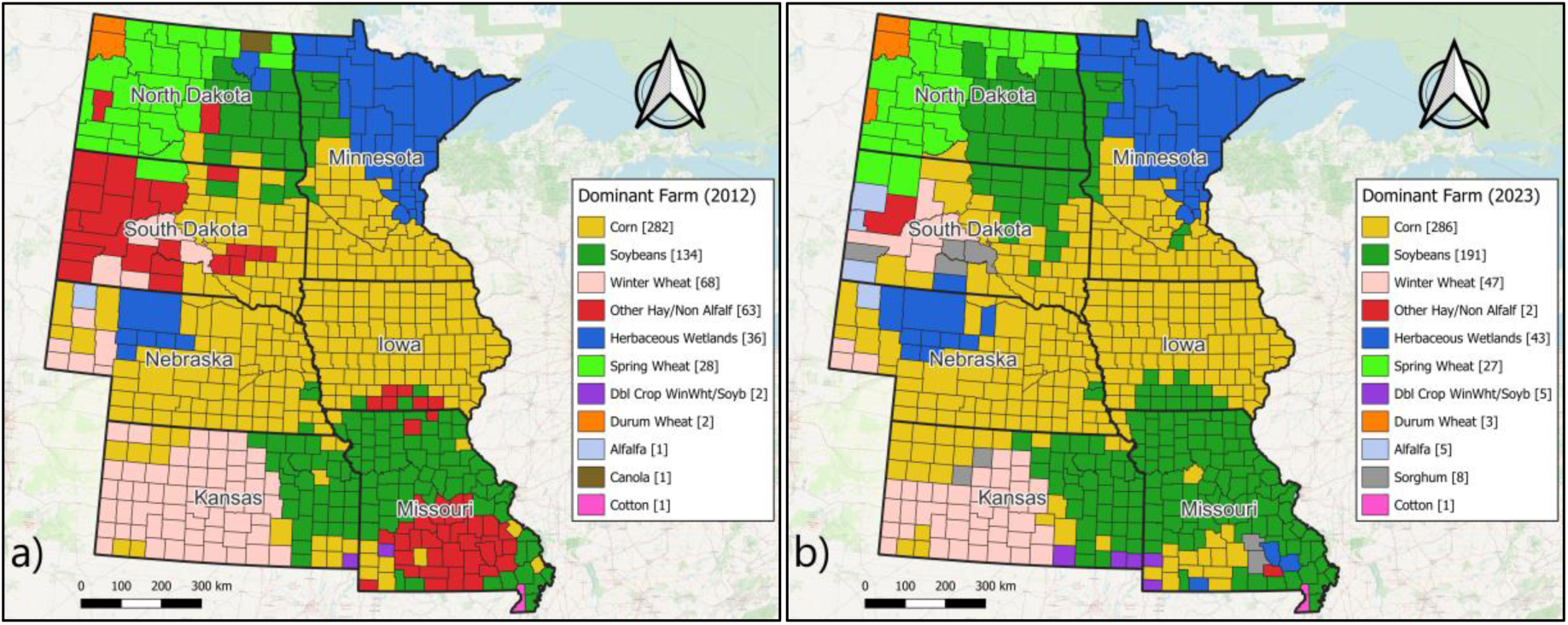
County level dominant agricultural characteristics of U.S Midwest region for a) 2012, and b) 2023.

The 2012 landscape was marked by a higher prevalence of “Other Hay/Non-Alfalfa” and a more distributed wheat presence, whereas the 2023 data reveal a significant consolidation into high-intensity Corn and Soybean production. Specifically, the number of counties where soybeans are the dominant crops rose from 134 in 2012 to 191 in 2023, reflecting an expansion of soybean acreage into the Dakotas and Missouri. This shift is critical from a safety perspective, as the transition from forage-based systems to mechanized row-cropping often correlates with increased exposure to high-capacity machinery and grain handling hazards (Ouattara et al., 2023).

As can be seen in Figure 1, The northern and western sections of the study area maintain distinct crop profiles that dictate seasonal risk cycles. North Dakota and northern Minnesota are defined by Spring Wheat and Herbaceous Wetlands, while Kansas and western Nebraska remain the primary corridors for Winter Wheat. The stability of the herbaceous wetland regions in northern Minnesota and the Dakotas, as seen in both Figure 1 (a) and (b), underscores the environmental constraints that influence farm size and the resulting spatial isolation of operators. Furthermore, the 2023 data highlight a sharp decline in counties dominated by “Other Hay” (dropping from 63 to 2), suggesting a transition toward more specialized crop outputs or changes in land management practices that favor row crops. These variations in Dominant Farm Type are not merely economic markers; they represent fundamental differences in the types of machinery used, the timing of peak labor demands, and the inherent occupational risks, such as tractor rollovers in hilly forage regions or grain entrapments in corn-heavy counties (Etienne et al., 2023; Jadhav et al., 2016).

### 2.2. Data Sources and Geospatial Preprocessing

This study utilizes a multi-layered geospatial framework to integrate longitudinal incident records with environmental and infrastructural data across the seven-state Midwest region. The primary dataset consists of agricultural incident records (2012–2023) sourced from the Central States Center for Agricultural Safety and Health (CS-CASH) surveillance program (CS-CASH, 2024). The study population focuses on self-employed farmers and ranchers, designated as operators. To ensure data integrity, these records were compiled into a cumulative master table and harmonized across key fields, including date, county, age, gender, severity outcome, and incident cause. Preprocessing involved cleaning duplicates and corrupted entries, alongside converting temporal data into a consistent date-time structure to facilitate seasonal and annual trend analysis. Administrative boundaries for the study area were constructed using a unified Midwest polygon and county-level subsets for Iowa, Minnesota, Nebraska, South Dakota, North Dakota, Kansas, and Missouri, providing the spatial foundation for all subsequent filtering and aggregation.

To contextualize these incidents within the agricultural landscape, USDA NASS Cropland Data Layer (CDL) raster wase integrated for each corresponding incident year. The CDL is a 30-meter resolution, geo-referenced, crop-specific land cover product primarily derived from satellite imagery, such as Landsat 8/9 and Sentinel-2, and validated using the Farm Service Agency (FSA) Common Land Unit (CLU) data (USDA NASS, 2024; Boryan et al., 2011). This integration ensures that the agricultural context, specifically the crop type, reflects the actual conditions at the time of the event, thereby accounting for the high degree of crop rotation and land-use changes inherent in Midwest (Yesilkoy and Demir, 2024). To address discontinuities in the raster data caused by non-farm areas or classification gaps, Euclidean Allocation was applied to the reclassified CDL surfaces. Euclidean Allocation is a spatial partitioning tool based on the Voronoi (Thiessen) polygon principle, which partitions a plane into regions close to each seed point (Guth & Klingel, 2012; Esri, 2024). In this study, it was used to bridge data gaps in the CDL raster by assigning the class of the nearest valid farm-type cell to null cells, creating a continuous surface that preserves the spatial structure of the production systems. Consequently, two farm-type descriptors were derived: Nearest Farm Type, assigned to precise incident locations, and County-Dominant Farm Type, used for county-level aggregates. Comparison between these descriptors showed a 30–35% match rate annually, suggesting they capture distinct but complementary aspects of the regional agricultural environment.

The spatial precision of incident records necessitated a dual-classification approach for geocoding to maintain analytical rigor. Location-known incidents, which include geographic coordinates or precise addresses, were utilized for high-resolution point-based analyses, such as kernel density estimation and proximity calculations. In contrast, County-only incidents, where only the county of occurrence was known, were assigned to the geographic centroid of the respective county. While approximately 4% of records (88 incidents) were excluded from spatial modeling due to insufficient data, they were retained for general descriptive statistics. Further data consistency checks resulted in the exclusion of a small number of records lacking critical temporal or demographic fields, though records missing only age (440 incidents) were retained as age serves as a secondary modifier rather than a structural requirement for spatial integration.

Finally, post-incident survivability was quantified through accessibility metrics for healthcare and communication infrastructure. EMS stations and hospital locations were sourced from the Homeland Infrastructure Foundation-Level Data (HIFLD) program and integrated into a road-network dataset (HIFLD, 2023). HIFLD is a collaborative effort between the Department of Homeland Security (DHS) and the Department of Defense (DoD) that provides a high-fidelity, unified database of critical infrastructure. For this study, the “EMS Stations” and “Hospitals” layers are critical for calculating travel time impedance in rural areas. Using ArcGIS Network Analyst, the study computed closest-facility measures to estimate the shortest-path travel distances and times for each location-known incident. A representative network-analysis output illustrating shortest-path connections and travel-time gradients is shown in Figure 2, where link colors indicate travel-time classes from incident locations to medical facilities.

**Figure 2:**
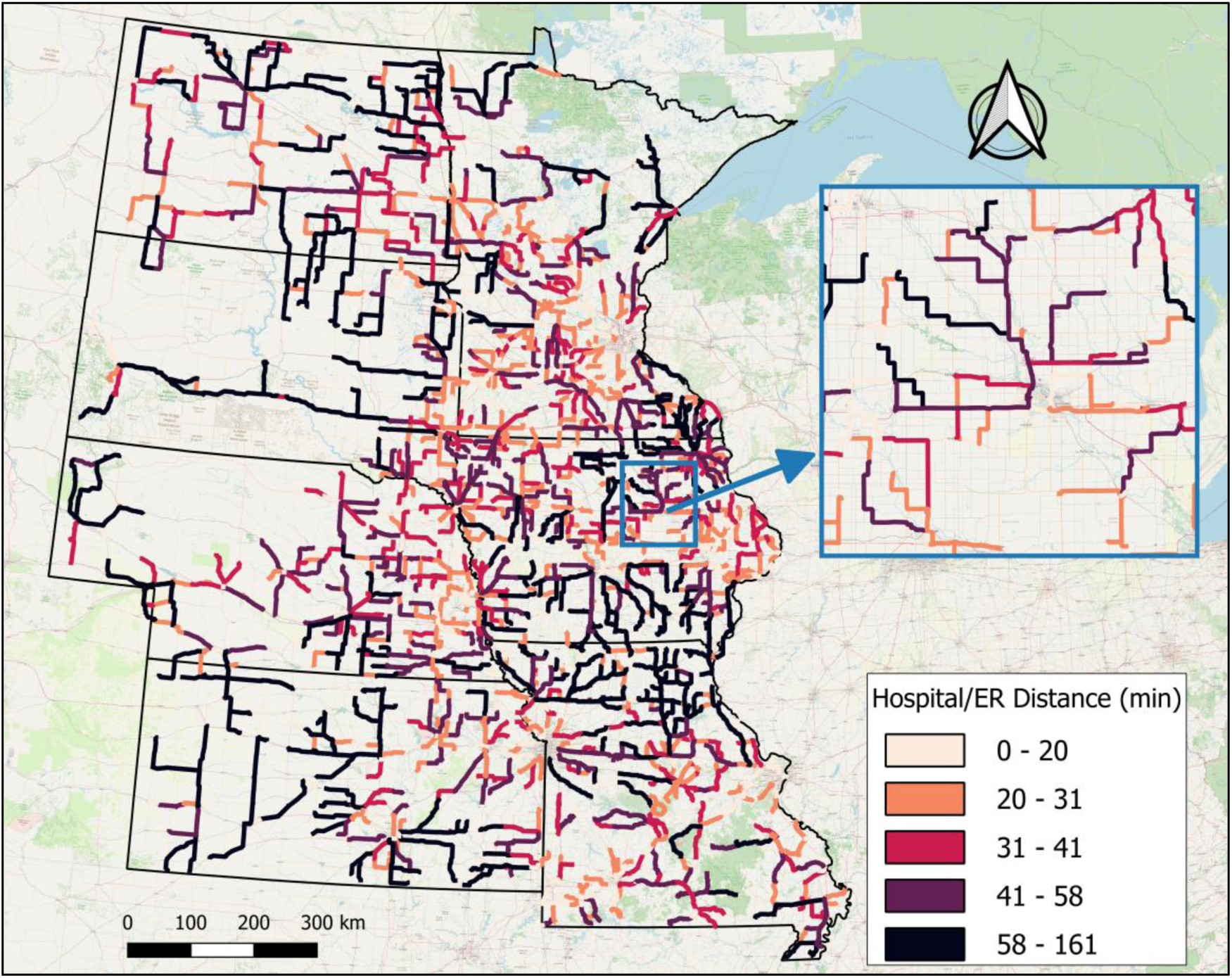
Network-based shortest-path connections from hospitals and emergency departments to incident locations across the Midwest.

Communication accessibility was proxied by calculating the Euclidean distance to the nearest cellular tower, utilizing FCC-maintained public access downloads (FCC, 2024). The Antenna Structure Registration (ASR) and Universal Licensing System (ULS) databases track the physical location and technical specifications of all registered cellular and radio towers in the U.S. In agricultural safety research, these locations serve as the primary proxy for cellular signal availability, which is the lifeline for lone workers needing to activate the emergency response chain after an incident. These metrics represent the structural constraints of the “golden hour”, the critical window in which prompt medical intervention significantly improves survival outcomes (Etienne et al., 2024a; Lerner & Moscati, 2001). All spatial layers were transformed into a consistent coordinate reference system and visually inspected to ensure alignment, providing a robust, standardized framework for the multi-criteria risk assessment.

### 2.3. Multi-Criteria Decision Making (MCDM)

The agricultural incident impact index is constructed using a deliberately structured set of parameters designed to represent the full incident lifecycle, from realized severity and intrinsic hazard through post-incident survivability to temporal relevance and agricultural context. This study employs a parameter set designed to remain compact and non-redundant while representing the dominant dimensions of agricultural incident impact. All parameter definitions, transformations, and weighting steps are explicitly documented to ensure methodological transparency and reproducibility.

#### 2.3.1. Parameter Selection and Normalization Hierarchical Structure

In spatial impact modeling, parameter selection requires balancing descriptive completeness with interpretability: too few parameters risk oversimplifying complex incident dynamics, while excessive or overlapping variables can dilute the influence of high-impact outcomes such as fatalities. Instead of treating all parameters as independent or equally influential, the selected variables are organized conceptually into functional subclasses that reflect their role in shaping incident impact. This structure provides transparency in parameter selection, clarifies representativeness across severity, access, time, and environment, and forms the conceptual basis for the subsequent weighting process.

The final parameter set used to construct the agricultural incident impact index consists of eight incident- and context-level variables organized to represent complementary dimensions of incident impact. All parameters are transformed into a unitless scale [0, 1], where larger values indicate a higher incident impact. Simple normalization is applied wherever possible, with composite formulations used only where necessary to encode meaningful constructions and provide robustness of the resulting index.

##### Fatality (FAT)

Fatality is the most direct and unambiguous indicator of realized severity and is central to agricultural injury surveillance and prevention prioritization (Penn State University, 2025). As death represents the maximum adverse outcome, this parameter anchors the upper bound of severity in the index. It is constructed using binary encoding as Equation 1:

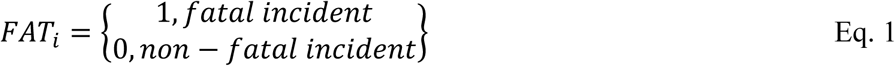

##### Incident Type (TYPE)

Incident type represents the intrinsic hazard associated with the underlying mechanism of injury, independent of the outcome of any single event. Mechanism has long been recognized as a key determinant of consequence in agricultural injury epidemiology, with machinery- and vehicle-related incidents contributing disproportionately to severe outcomes (Jadhav et al., 2015). Tractors and heavy equipment are consistently identified as dominant sources of injury, particularly through rollovers, runovers, and entanglement pathways (Qi et al., 2024), and tractor overturns remain a persistent cause of agricultural fatalities across regions and time periods (Myers and Hendricks, 2010). Grain bin entrapments exhibit similarly high fatality proportions due to confined-space conditions and rescue difficulty (Issa et al., 2016), while all-terrain vehicle incidents account for a substantial share of occupational vehicle deaths, frequently involving overturn mechanisms (Helmkamp et al., 2010).

Collectively, this evidence indicates that risk is distributed across multiple high-consequence mechanisms rather than dominated by a single incident category. To reflect both the inherent danger of each mechanism and its relative occurrence, incident type is modeled as a composite measure that integrates two components: the fatality rate associated with each type and its share of total incidents. Each component is normalized to the range [0, 1] and combined with equal influence. The parameter is a composite measure integrating fatality rate (FR) and the share of total incidents (SH) to avoid over-emphasizing rare events, by using Equation 2-4:

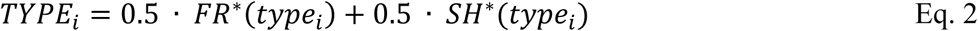

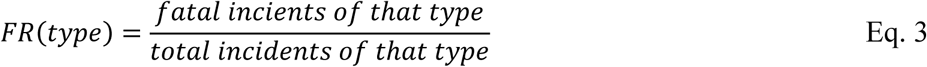

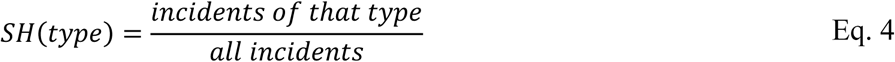

where (·) denotes normalization to [0,1]. This formulation prevents rare but highly lethal mechanisms from being overstated while also avoiding dominance by common but lower-consequence events, allowing the parameter to capture intrinsic mechanism hazard in an epidemiologically grounded and balanced manner.

##### Temporal Relevance (REC)

Research in dynamic risk modeling treats recent data as more relevant for outcome forecasts (Bull et al., 2020). This parameter reflects evolving agricultural practices and safety interventions. More recent incidents are treated as more representative of current risk conditions. The incident year is normalized across the 2012–2023 analysis window, with more recent incidents receiving higher values.

##### People Involved (PEOP)

Incidents involving multiple individuals represent greater immediate impact and higher potential for straining emergency response capacity (Alpert and Kohn, 2023). This is treated as a moderate impact modifier instead of a primary driver, because multi-person incidents are relatively rare and do not define the intrinsic hazard mechanism.

##### Age (AGE)

Included as a vulnerability modifier, as older workers and very young individuals are more susceptible to severe outcomes (Smith and Pegula, 2020). Due to potential noise and inconsistent reporting at the spatial incident level, it is assigned a small relative influence.

##### EMS/Hospital Closeness (EM)

It represents accessibility to emergency medical care following an incident. Emergency response and transport time are strongly associated with survival outcomes in rural trauma settings (Alruwaili and Alanazy, 2022; Miller et al., 2020). Rural EMS runs are consistently longer than urban ones, reflecting structural inequities in healthcare access (Byrne et al.,2019). Network-based travel time from each incident to the nearest EMS station and/or hospital is computed using road network analysis and normalized to [0,1], with larger values indicating poorer access (longer response/transport).

##### Tower Closeness (TOW)

Proximity to cellular infrastructure serves as a proxy for the ability to activate and coordinate emergency response. Limitations in communication infrastructure, such as unreliable or insufficient cellular connectivity, can hinder EMS activation, delay medical direction, and reduce the effectiveness of emergency response coordination, particularly in rural and remote settings (Karra et al., 2024). Euclidean distance to the nearest tower is used as a relative indicator.

##### Farm-Type Fatality Ratio (FARM)

This encodes the lethality of the surrounding production environment. Literature suggests different systems (e.g., livestock vs. crops) have distinct injury patterns (Outtara et al., 2023). In methodology manner, crop-specific proximity comparisons (e.g., within 500 m of a specific crop vs not)—another direct example of treating crop type near a location as the exposure/context variable (Simões et al., 2022). For each farm type, the fatality ratio is computed by Equation 5 where it functions as a contextual modifier with lower influence due to classification uncertainty.

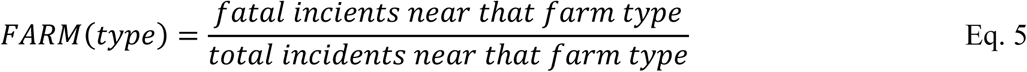

#### 2.3.2. Analytic Hierarchy Process (AHP)

The Analytic Hierarchy Process (AHP) was used to convert the eight incident-impact criteria into a consistent set of weights that represent their relative contribution to overall incident impact (Saaty, 1987). AHP pairwise judgments on a ratio scale (Saaty’s 1–9 AHP scale; Table 1) and derives a weight vector that summarizes these comparisons.

**Table 1.** AHP and FAHP Scales for Pairwise Comparison Matrix (Putra et al., 2018).

| Parameters | AHP Scale | TFN Scale | Reciprocal TFN Scale |
| --- | --- | --- | --- |
| Equally Important | 1 | (1, 1, 1) | (1, 1, 1) |
| Equal to Moderate | 2 | (0.5, 1, 1.5) | (0.67, 1, 2) |
| Moderately Important | 3 | (1, 1.5, 2) | (0.5, 0.67, 1) |
| Moderate to Important | 4 | (1.5, 2, 2.5) | (0.4, 0.5, 0.67) |
| Important | 5 | (2, 2.5, 3) | (0.33, 0.4, 0.5) |
| Important to High Important | 6 | (2.5, 3, 3.5) | (0.28, 0.33, 0.4) |
| High Important | 7 | (3, 3.5, 4) | (0.25, 0.28, 0.33) |
| High to Extreme Important | 8 | (3.5, 4, 4.5) | (0.22, 0.25, 0.28) |
| Extremely Important | 9 | (4, 4.5, 4.5) | (0.22, 0.22, 0.25) |

##### Ordinal Hierarchy Building

Pairwise judgments were constrained to preserve ordinal consistency with observed severity patterns in agricultural injury data that parameters representing direct harm and intrinsic hazard were consistently prioritized over contextual modifiers. Internal coherence of these judgments was subsequently evaluated through systematic consistency checks. Within this framework, FAT is assigned the highest importance, followed by TYPE, reflecting that fatal outcomes and high-lethality mechanisms constitute the most direct expressions of agricultural incident impact. Parameters associated with post-incident conditions, EM, and TOW, are ranked below severity-related parameters, as they condition survivability after an incident has occurred but do not define the intrinsic magnitude of harm.

REC is positioned below survivability parameters to capture temporal relevance without allowing time effects to outweigh severity or access-related constraints. Parameters that function primarily as modifiers, such as PEOP, FARM, and AGE are intentionally assigned to lower influence. PEOP amplifies event-level impact without representing an intrinsic hazard mechanism, FARM provides agricultural context with recognized classification uncertainty and partial overlap with mechanism information, and AGE serves as a vulnerability proxy that is spatially noisy at the incident level. This prioritization allows severity- and mechanism-driven signals to dominate the index, followed by survivability and temporal relevance, while contextual and demographic factors function as secondary modifiers that refine interpretation without distorting the overall spatial impact pattern.

##### Pairwise Comparison Matrix

The comparative matrix represents the relative importance of numerical values based on the AHP scale. The expression in Equation 6 issued for constructing a pairwise matrix:

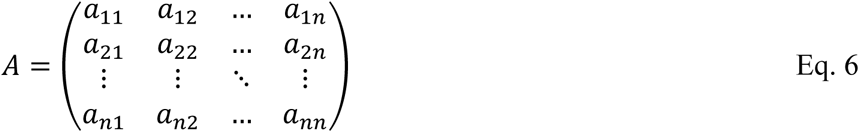

where *a*_*ij*_ is the importance ratio of criterion *i* relative to criterion *j*, *n* is the number of criteria (here *n* = 8). When the criterion in the row is judged more important than the criterion in the column, *a*_*ij*_ is assigned a value from 1 to 9 according to the AHP scale in Table 1. Conversely, when the row criterion is less important, the reciprocal value 1/*a*_*ij*_is assigned, producing a positive reciprocal matrix.

##### Relative Weights

After building the pairwise matrix, the column normalization is required by applying the following Equation 7 to represent each parameter’s weight proportionally:

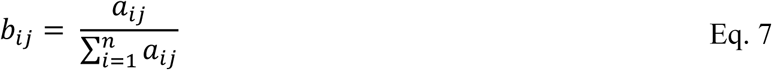

where *b*_*ij*_ is the normalized entry and the denominator is the sum of column *j*. The normalized matrix is then used to derive criterion weights. Row means of the normalized matrix provide the AHP weights, which are calculated by Equation 8:

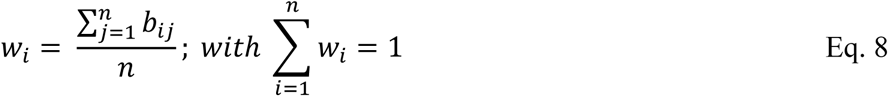

where *w*_*i*_ is the weight of criterion *i*. This normalization expresses each criterion as a proportional share of the overall comparison structure, allowing the weights to be directly comparable.

##### Consistency Validation

The Consistency Ratio (CR) of the pairwise judgments are calculated by comparing the Consistency Index (CI) to the corresponding Random Index (RI) by using Equation 9:

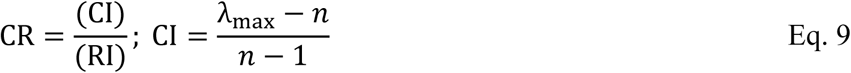

where λ_max_ is the principal (i.e., largest) eigenvalue from the pairwise matrix, and *n* is the number of parameters (Malczewski, 1999). As can be seen in Table 2, RI is random index and n is our dependent count, which is 8. A matrix is considered consistent when CR<0.10 (Saaty, 1987). As shown in Table 2, the corresponding RI value is selected based on the matrix size (*n* = 8). Table 3 presents the finalized pairwise comparison matrix together with the corresponding consistency validation results, which form the basis for the subsequent fuzzy analysis.

**Table 2.** Random Index (RI) values (Saaty, 1987)

| **n** | 1 | 2 | 3 | 4 | 5 | 6 | 7 | **8** | 9 |
| --- | --- | --- | --- | --- | --- | --- | --- | --- | --- |
| **RI** | 0 | 0 | 0.58 | 0.90 | 1.12 | 1.24 | 1.32 | **1.41** | 1.45 |

**Table 3.** Pairwise Comparison Matrix for all the parameters.

| Parameters | FAT | TYPE | REC | PEOP | AGE | EM | TOW | FARM |
| --- | --- | --- | --- | --- | --- | --- | --- | --- |
| **FAT** | 1 | 3 | 5 | 7 | 7 | 3 | 7 | 7 |
| **TYPE** | 0.33 | 1 | 3 | 7 | 7 | 2 | 5 | 7 |
| **REC** | 0.2 | 0.33 | 1 | 3 | 7 | 0.33 | 1 | 5 |
| **PEOP** | 0.14 | 0.14 | 0.33 | 1 | 3 | 0.2 | 0.5 | 2 |
| **AGE** | 0.14 | 0.14 | 0.14 | 0.33 | 1 | 0.14 | 0.2 | 0.5 |
| **EM** | 0.33 | 0.5 | 3 | 5 | 7 | 1 | 3 | 7 |
| **TOW** | 0.14 | 0.2 | 1 | 2 | 5 | 0.33 | 1 | 3 |
| **FARM** | 0.14 | 0.14 | 0.2 | 0.5 | 2 | 0.14 | 0.33 | 1 |
| $\lambda_{\max} = 8.54$; $CI=0.77$; $RI=1.41$; $CR=0.05 < 0.1$ | | | | | | | | |

As shown in Table 3, the pairwise comparison values were constructed using the AHP scale and validated using the consistency measures defined in Equations (9) and (10). The resulting Consistency Ratio (CR=0.05) is below the acceptable threshold of 0.10 so the judgments are internally consistent and suitable for subsequent analysis. However, AHP remains sensitive to shifts in criteria and subjective judgments, which may introduce additional uncertainty into the weighting process (Pourghasemi et al., 2012). To better capture this uncertainty in pairwise evaluations, the analysis is extended using the Buckley Fuzzy AHP approach.

### 2.3.3. Geometric Fuzzy Analytic Hierarchy Process (Geometric-FAHP)

Classical AHP requires decision-makers to express each pairwise comparison as a single exact ratio. In practice, those judgments are often stated in linguistic terms (e.g., “important”, “very important”), and their numerical representation may vary slightly across evaluators. To capture this uncertainty without changing the preference structure already validated through AHP consistency, this study applies a fuzzy extension of AHP based on triangular fuzzy numbers (TFNs) (van Laarhoven & Pedrycz, 1983). The FAHP implementation adopts the fuzzy geometric mean method proposed by Buckley (1985).

The main reason for using Buckley’s approach is that it keeps the same multiplicative logic as ratio-based AHP but allows each comparison to carry a bounded uncertainty range. In other words, it reduces the “dominance” behavior of minimum-based possibility comparisons of fuzzy AHP. In such approaches, repeatedly lower judgments can drive some criteria toward disproportionately small weights and allow highly dominant factors to absorb the relative contribution of lower-priority criteria. For an impact index in which severity-related criteria (e.g., fatality and incident type) are expected to remain prominent while contextual modifiers retain influence, Buckley’s method yields stable and interpretable weight estimates.

#### Fuzzification of pairs

The crisp comparison matrix *A* is converted into a fuzzy reciprocal matrix *Ã* using Equation 10 where *l*_*ij*_ < *m*_*ij*_ < *u*_*ij*_ are the lower, most likely (modal), and upper bounds of the judgment that criterion *i* is more important than criterion *j*. Reciprocal comparisons are assigned using the reciprocal TFN scale (Table 1), so *ã*_*ij*_ maintains linguistic reciprocity within the matrix.

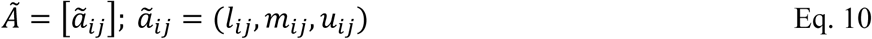

#### Fuzzy geometric mean aggregation

For each criterion *i*, the fuzzy geometric mean *g̃*_*i*_ is computed by aggregating the TFNs across the corresponding row using Equation 11 where *n* is the number of criteria, and *g̃*_*i*_ is the fuzzy geometric mean of criterion *i*. The exponent 1/*n* (the n^th^ root) converts the cumulative product into a geometric mean, preventing scale inflation as the number of criteria increases and maintaining consistency with ratio-based aggregation in AHP (Buckley, 1985). In implementation, the TFN multiplication is performed component-wise (lower with lower, modal with modal, upper with upper), and the n^th^ root is applied to each component. This yields our geometric mean *g̃*_*i*_ as a triangular fuzzy number that preserves uncertainty bounds while synthesizing pairwise judgments for criterion *i*.

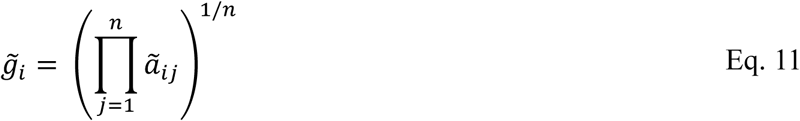

#### Normalization to fuzzy weights

After computing the fuzzy geometric mean, these values must be normalized to obtain comparable fuzzy weights. The total fuzzy sum of geometric means is defined in Equation 12:

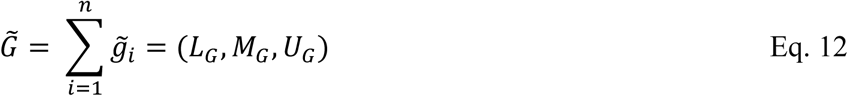

where *L*_*G*_, *M*_*G*_, *U*_*G*_ represents the lower, modal, and upper components of the aggregated TFN. To normalize, each geometric mean *g̃*_*i*_ is multiplied by the inverse *G̃*, which is *G̃*^−1^, to preserve the triangular ordering *l* < *m* < *u*. The normalized fuzzy weight for criterion *i* is then obtained using Equation 13:

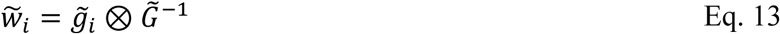

where ⊗ is component-wise multiplication. Through this operation, the fuzzy geometric means are proportionally rescaled so that all criteria are expressed relative to the total comparison structure, providing normalized fuzzy weights suitable for subsequent defuzzification.

#### Defuzzification and crisp weights

Finally, to obtain a single weight per criterion for the incident impact index, the fuzzy weights are defuzzified and normalized using the centroid (center-of-area) method as shown in Equation 14:

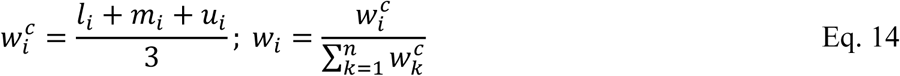

where the term 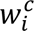 represents the defuzzified (crisp) weight obtained. In the normalization, *n* is the total number of criteria and *k* is the summation index across the criteria set. The resulting *w*_*i*_ values are the final normalized weights generated by the Buckley FAHP procedure and are subsequently used in the weighted aggregation of the normalized incident parameters to construct the agricultural incident impact index (Equation 15).

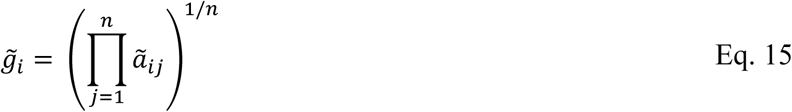

The fuzzy geometric means are then normalized and defuzzied using the centroid (center-of-area) method to obtain the final crisp weights (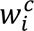) used for the index (Equation 16):

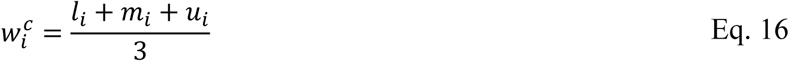

This ordering ensures that severity- and mechanism-driven signals (FAT and TYPE) dominate the index, followed by survivability (EM and TOW), while contextual effects (PEOP, FARM, and AGE) refine the interpretation without distorting spatial impact patterns.

### 2.3.4. Visualization Products and Spatial Smoothing

Visualization is incorporated as a methodological component to examine spatial and temporal structure in agricultural incident data and to communicate patterns prior to and alongside index development. Graphical summaries are used to evaluate annual trends, demographic concentration, and incident-type variability, providing contextual insight before formal weighting and aggregation. Histograms of AHP- and Geometric-FAHP–derived weights are additionally used to assess how relative importance is distributed across criteria and to observe how the fuzzy formulation moderates dominance compared to the crisp AHP solution. Spatial representation at the county level aligns the analysis with administrative units commonly used in emergency response, occupational safety planning, and public health decision-making. County-based mapping allows regional contrasts and structural disparities in agricultural injury risk to be interpreted within a policy-relevant framework.

To complement areal representations, Kernel Density Estimation (KDE) is applied to convert point-based incident locations into continuous spatial intensity surfaces. KDE identifies localized clustering and hotspot patterns without imposing administrative boundaries. The resulting density landscape highlights overlooked spatial concentrations, supports exploratory scenario assessment, and provides an intuitive overview of incident patterns for planning and decision support (Duran et al., 2025b). The estimated density at spatial location is expressed using Equation 17:

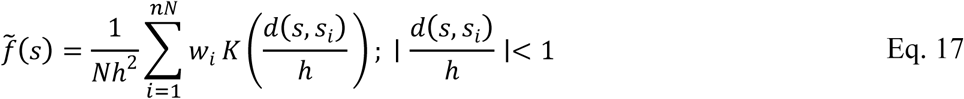

where *N* is the number of incidents, *s*_*i*_ is the location of incident *i*, *d*(*s*, *s*_*i*_) is the distance between the evaluation location and the incident, ℎ is the bandwidth controlling the smoothing scale, *w*_*i*_ represents the weight assigned to incident, and *K*(⋅) is the kernel function (Silverman, 1986; Diggle, 1985, 1990). The condition ∣ *d*(*s*, *s*_*i*_)/ℎ ∣< 1 indicates that only incidents within the specified bandwidth contribute to the density estimate. The bandwidth determines the spatial scale of influence, where larger values emphasize regional trends, and smaller values preserve localized variation. Building on this compact-support condition, a quartic (biweight) kernel is adopted due to its smooth distance decay while restricting influence on the defined neighborhood. This balance allows local hotspots to be identified without excessive smoothing or artificial discontinuities and is well suited for exploratory spatial analysis of point-based incident data. Both unweighted KDE surfaces (incident frequency) and weighted KDE surfaces are generated. In the weighted formulation, incident-level impact scores derived from AHP and Geometric-FAHP are used as *w*_*i*_. This shifts interpretation from spatial concentration of events to concentration of higher-impact incidents.

## 3. Results and Discussion

This section examines the empirical patterns of agricultural incidents across the study region and evaluates how differences in frequency, severity, and spatial distribution contribute to overall risk. Descriptive statistics, spatial mapping, and index-based weighting results are used together to distinguish exposure-driven activity patterns from severity-driven impact concentrations and to assess how the AHP and Geometric-FAHP frameworks modify the regional representation of agricultural incident risk.

### 3.1. General Incident Characteristics

The overall characteristics of the incident records were first examined to establish a baseline understanding of incident frequency and mechanisms most contributing to fatal outcomes. Tables 4 summarizes state-by-state distribution of fatal (F) and injury-causing (I) incidents by incident type across the Midwest study region. Incident counts are strongly concentrated in Iowa, Minnesota, and Missouri, with Iowa alone reporting 259 fatal and 266 injury cases. This distribution largely reflects agricultural exposure; these states contain highly intensive row-crops and mixed farming systems with higher machinery utilization and operator hours. Consequently, raw totals primarily indicate where agricultural activity is most concentrated rather than where conditions are inherently more dangerous.

A clearer distinction appears when analyzing incident mechanisms. Fatal incidents are disproportionately associated with tractors, grain bins, ATV/UTV vehicles, and other heavy equipment categories. These mechanisms are characterized by high energy transfer, rollover potential, or confined-space entrapment, which significantly reduces survivability. Their prominence in fatal incidents indicates that a relatively small set of high-consequence mechanisms governs most loss-of-life outcomes. By contrast, injury-causing incidents are dominated by roadway and entanglement events. This distinction between frequent but survivable incidents and less frequent yet high consequence incidents indicates that incident counts alone cannot fully reflect overall impact.

**Table 4.** State-by-State distribution of Fatal (F) and Injury-Causing (I) agricultural incidents by incident type.

| Incident Type | Iowa |  | Kansas |  | Minnesota |  | Missouri |  | Nebraska |  | N. Dakota |  | S. Dakota |  |
| --- | --- | --- | --- | --- | --- | --- | --- | --- | --- | --- | --- | --- | --- | --- |
|  | F | I | F | I | F | I | F | I | F | I | F | I | F | I |
| Tractor | 67 | 50 | 34 | 19 | 51 | 50 | 83 | 61 | 33 | 23 | 17 | 7 | 12 | 2 |
| Roadway | 38 | 116 | 19 | 43 | 25 | 73 | 27 | 76 | 31 | 47 | 12 | 22 | 4 | 12 |
| Livestock | 7 | 3 | 4 | 3 | 6 | 1 | 9 | 6 | 4 | 9 | 2 | 2 | 1 | 2 |
| ATV/UTV | 27 | 8 | 7 | 5 | 9 | 8 | 15 | 16 | 16 | 15 | 9 | 3 | 10 | 4 |
| Skid-Loader | 9 | 5 | 3 | 0 | 14 | 4 | 0 | 0 | 0 | 0 | 0 | 0 | 4 | 0 |
| Grain Bin | 26 | 19 | 3 | 7 | 18 | 15 | 7 | 4 | 16 | 11 | 9 | 0 | 5 | 3 |
| Entanglement | 8 | 18 | 0 | 3 | 9 | 44 | 4 | 6 | 7 | 7 | 5 | 13 | 3 | 4 |
| Other | 77 | 47 | 46 | 16 | 61 | 43 | 40 | 22 | 46 | 35 | 29 | 19 | 17 | 13 |
| State Total | 259 | 266 | 116 | 96 | 193 | 238 | 185 | 191 | 153 | 147 | 83 | 66 | 56 | 40 |

To further quantify these differences, the fatality rate for each incident type (shown in Figure 3) was calculated by dividing the total number of fatal incidents by the total number of incidents (fatal plus injury) within that category. Regional data indicates that equipment-related categories remain above the average fatality rate of around 50%, with skid-loader incidents emerging as the most lethal mechanism at 76.9%. High fatality proportions are also noted for tractors (58.3%), grain bins (58.7%), and ATV/UTV events (61.2%). In contrast, roadway (28.6%) and entanglement (27.5%) incidents fall at the lower end of the lethality range. While frequent, these events are more survivable than those involving heavy machinery or confined spaces. This pattern points to two distinct risk structures within agricultural safety: machinery incidents tend to be less frequent but more severe, whereas roadway and entanglement incidents are more common but less likely to result in a fatal outcome. Because the likelihood of fatal outcome varies notably by mechanism, treating all incident types equally would obscure important differences in impact. These findings support incorporating mechanism-based severity weighting into the impact index so that the assessment reflects both occurrence and consequence.

**Figure 3.**
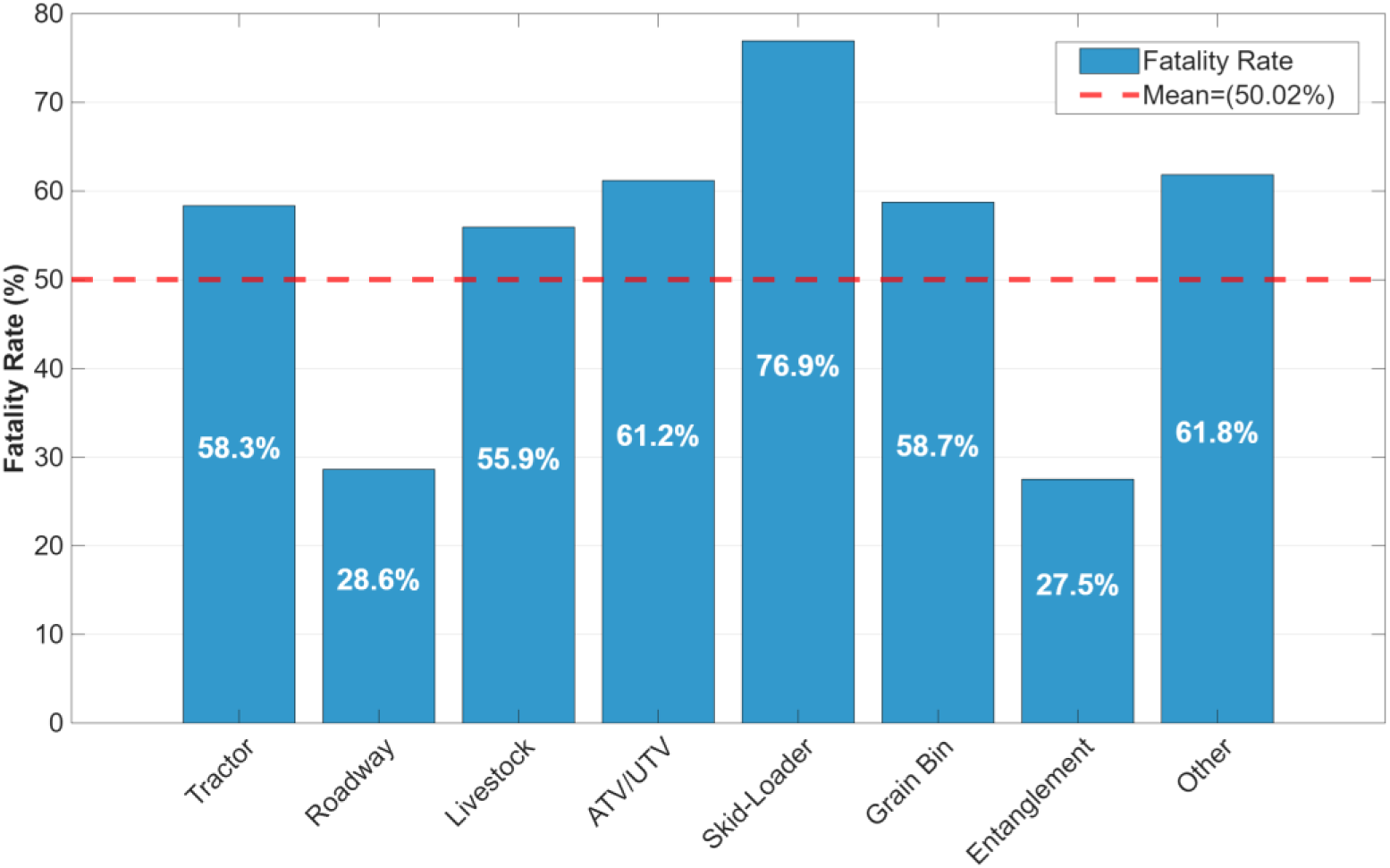
Incident-type-specific fatality rates for agricultural incidents across the Midwest region.

State-level fatality proportions further reinforce these findings. Although Iowa reports the larger number of incidents, only about 49% of its incidents are fatal, which is comparable to Missouri and lower than several other states. In contrast, South Dakota, North Dakota, and Kansas show higher fatality proportions of 58%, 56%, and 55%, respectively, despite having fewer total incidents. Minnesota exhibits the lowest proportion at 45%. Frequency and impact therefore represent separate dimensions of risk and evaluating either measure in isolation provides an incomplete picture of agricultural safety conditions.

Demographic patterns further clarify the distribution of agricultural risk. Incident records are heavily male dominated, with males representing 78% of all cases and 85% of fatal outcomes, indicating that exposure to hazardous agricultural activities remains heavily concentrated among male workers. Although gender was not incorporated into the impact scoring framework, this imbalance reflects underlying differences in task exposure and workforce composition within agricultural operations. Age patterns show a shift toward older cohorts. Although incidents are recorded across all age groups, total case accumulation becomes more pronounced among individuals aged roughly 50-80 years. More importantly, the fatality rate itself increases with advancing age, indicating that older individuals face a higher probability of death once an incident occurs. For instance, the 60 to 70 age group accounts for 314 incidents and 204 fatalities, representing the highest fatal burden in the dataset. Younger groups, by contrast, experience comparatively lower fatal shares relative to their total incidents. These findings indicate that agricultural risk is driven by a combination of task exposure and age-related vulnerability, particularly among older male workers.

### 3.2. Geospatial Clustering and Localized Hotspots

While state-level summaries quantify overall counts and outcomes, they do not reveal the precise geographic distribution of agricultural risk. Spatial patterns are critical because areas with similar incident totals can experience different outcomes depending on the localized concentration of severe incident mechanisms. By mapping total, injury-causing, and fatal incidents at the county level, it is possible to distinguish between exposure-driven activity clusters and high-lethality hotspots.

As illustrated in Figure 4, the total incident map identifies several pronounced exposure clusters. The most significant concentrations appear in central Minnesota, specifically in Stearns and Morrison counties, and in northwestern Iowa, notably in Sioux and Plymouth counties. Secondary clusters are visible in parts of Missouri and central Kansas. This spatial footprint largely mirrors the intensity of regional agricultural operations, where higher levels of farm activity and machinery usage naturally correlate with higher reporting volumes. Consequently, the total incident surface serves as a baseline indicator of agricultural exposure. In contrast, injury-causing incidents exhibit noticeably smoother spatial distribution.

**Figure 4.**
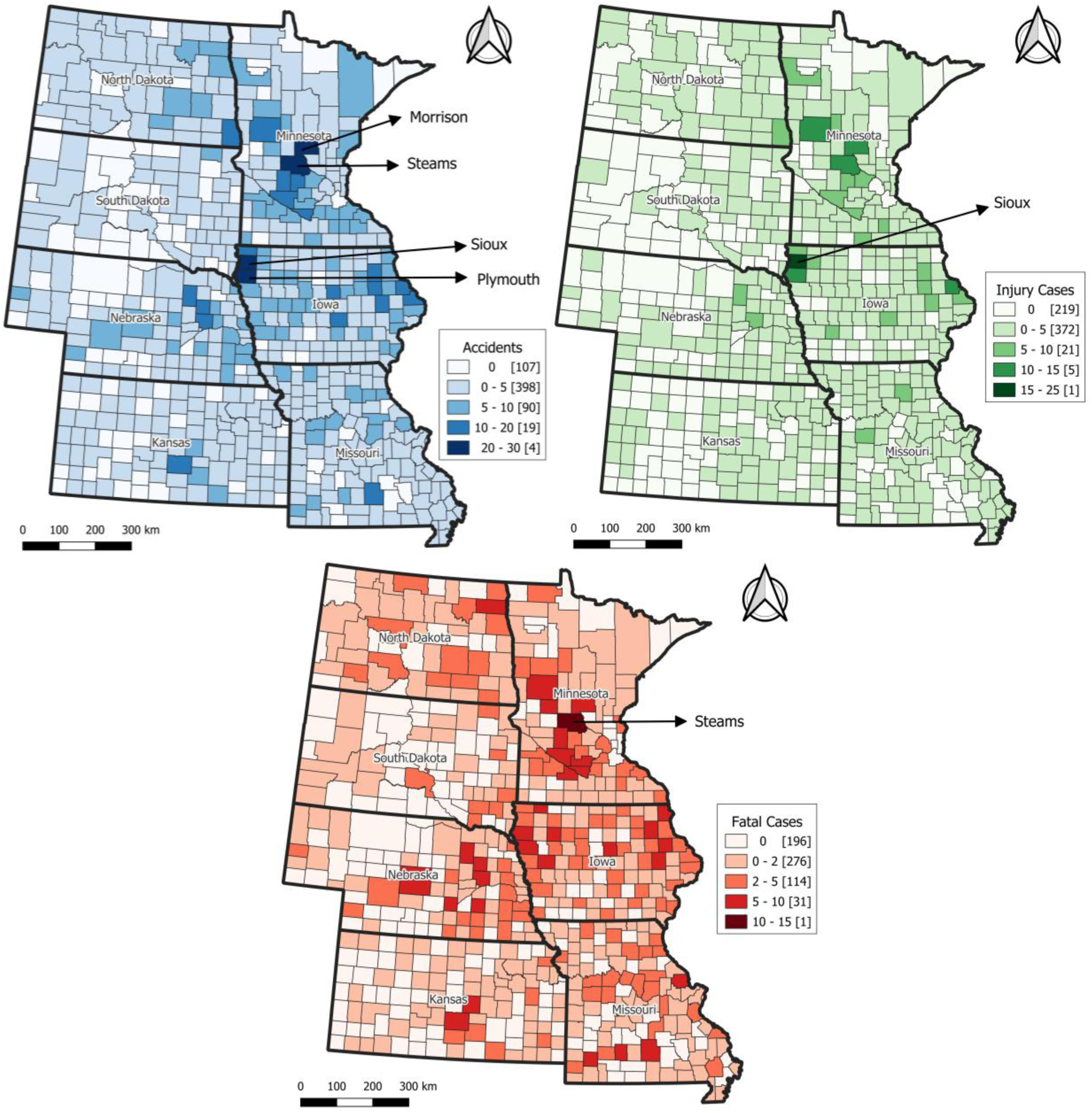
County-level distribution of total, injury-causing, and fatal agricultural incidents in the Midwest region.

Although Sioux County, Iowa remains a prominent hot spot, most counties show moderate values without sharp localized peaks. This broader distribution aligns with the prevalence of roadway and entanglement incidents in the injury dataset. Because these mechanisms occur more routinely across the agricultural landscape and are more survivable, their spatial footprint tracks general agricultural activity rather than forming isolated, high-intensity hotspots.

Fatal incidents, however, present a distinct and more fragmented spatial structure. The most prominent hotspot occurs in Stearns County, Minnesota, which stands out clearly from its surrounding areas. Additional discrete clusters appear in part of Iowa, southern Missouri, central Kansas, eastern Nebraska, and eastern North Dakota. Compared to total and injury patterns, fatalities are concentrated in fewer counties and form sharper, more isolated peaks. This divergence indicates that fatal outcomes are not simply proportional to incident frequency; instead, they are heavily influenced by the presence of high-severity mechanisms such as heavy machinery, grain handling, rollover events, and confined-space incidents.

The contrast among these three maps demonstrates that frequency and impact do not share identical spatial behavior. If severity played a negligible role, the fatal distribution would closely mirror the total incident pattern. Instead, the observed divergence proves that relying solely on raw incident counts would significantly underrepresent localized high-consequence areas. These findings provide the methodological bridge to the subsequent impact index analysis, where multi-criteria weighting is used to capture counties where agricultural incidents are both frequent and inherently more dangerous.

### 3.3. Multi-Criteria Weighting Analysis and Score Distribution

To complement the descriptive and spatial patterns, a set of impact indices was generated to integrate incident frequency, outcome severity, and contextual vulnerability into singular quantitative measures. Instead of evaluating incidents only by occurrence or location, the indices assign relative importance to fatal consequences, incident mechanisms, emergency accessibility, and supporting demographic and environmental factors. The weighting schemes derived from AHP and Geometric-FAHP therefore provide two complementary views of impact: one emphasizing severity-driven contrasts and the other capturing broader and more gradual vulnerability patterns.

As shown in Table 5, the AHP results prioritize direct outcome impact, with Fatality (FAT) and Incident Type (TYPE) accounting for nearly 60% of the total weight. This is followed by emergency medical accessibility (EM), reflecting a focus on the most catastrophic events. In contrast, Geometric-Fuzzy AHP reduces the dominance of these top criteria and redistributes weight toward contextual and exposure-related factors, such as Recency (REC), Tower Closeness (TPW), and Farm-Type (FARM), while maintaining a similar emphasis on medical access. This shift reflects the uncertainty-aware nature of Geometric-FAHP, producing a more balanced weighting structure that moderates extreme hotspots and increases sensitivity to accessibility and contextual vulnerability compared to crisp AHP.

**Table 5.**
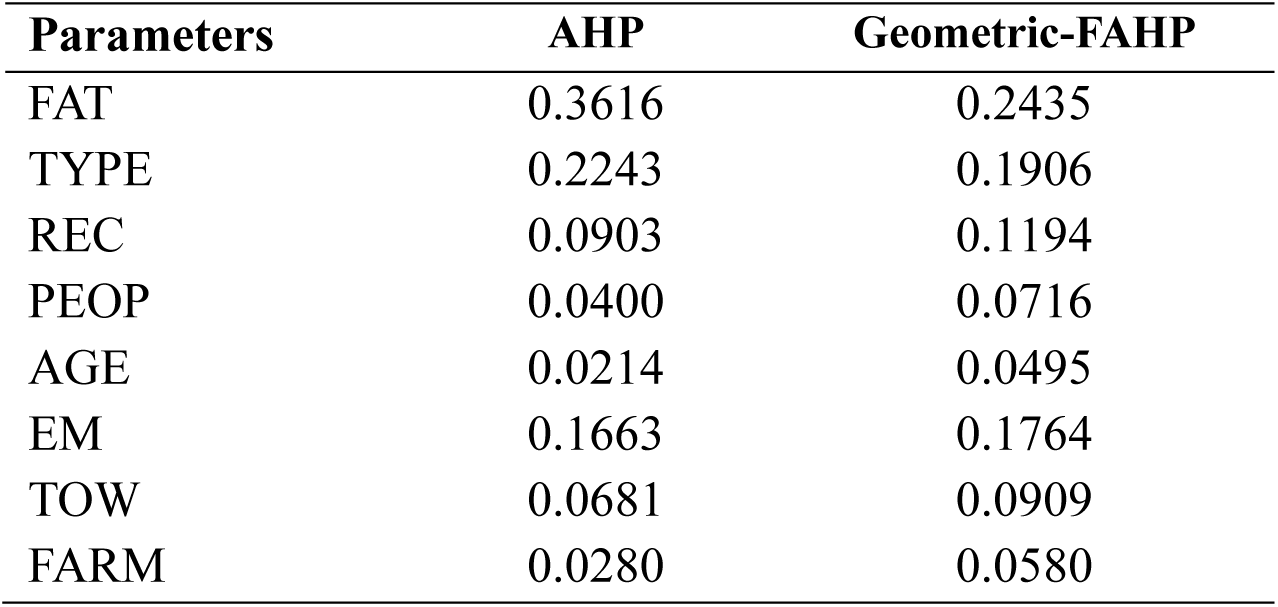
Criterion weights derived from AHP and Geometric-FAHP impact index framework.

| Parameters | AHP | Geometric-FAHP |
| --- | --- | --- |
| FAT | 0.3616 | 0.2435 |
| TYPE | 0.2243 | 0.1906 |
| REC | 0.0903 | 0.1194 |
| PEOP | 0.0400 | 0.0716 |
| AGE | 0.0214 | 0.0495 |
| EM | 0.1663 | 0.1764 |
| TOW | 0.0681 | 0.0909 |
| FARM | 0.0280 | 0.0580 |

Instead of treating AHP and Geometric-FAHP as competing approaches, the two weighting schemes were used in a complementary manner. The AHP framework concentrates weight on outcome severity (FAT and TYPE), producing greater contrast and emphasizing localized, high-impact hotspots where catastrophic incidents dominate. In contrast, Geometric-FAHP distributes weights more evenly across parameters by incorporating uncertainty in pairwise comparisons, thereby increasing the influence of contextual and accessibility-related factors such as emergency response and recency. Consequently, Geometric-FAHP highlights broader regional vulnerability patterns that may not be captured by severity alone.

Taken together, the two metrics provide a dual perspective, enabling the identification of both sharp local peaks and more diffuse, system-wide risk structures and thereby supporting a more comprehensive assessment of agricultural incident impacts. To illustrate these differences, Figure 5 presents a histogram of case-level impact scores for both AHP and Geometric-FAHP using identical interval bins that enable direct comparison of their distributions and overlaps. Geometric-FAHP results form a unimodal and centrally concentrated pattern, with the majority of incidents clustered within the mid-range of 0.45 to 0.65. This concentration reflects the tendency of the fuzzy structure to moderate extreme values and produce a more continuous gradation of impact.

**Figure 5.**
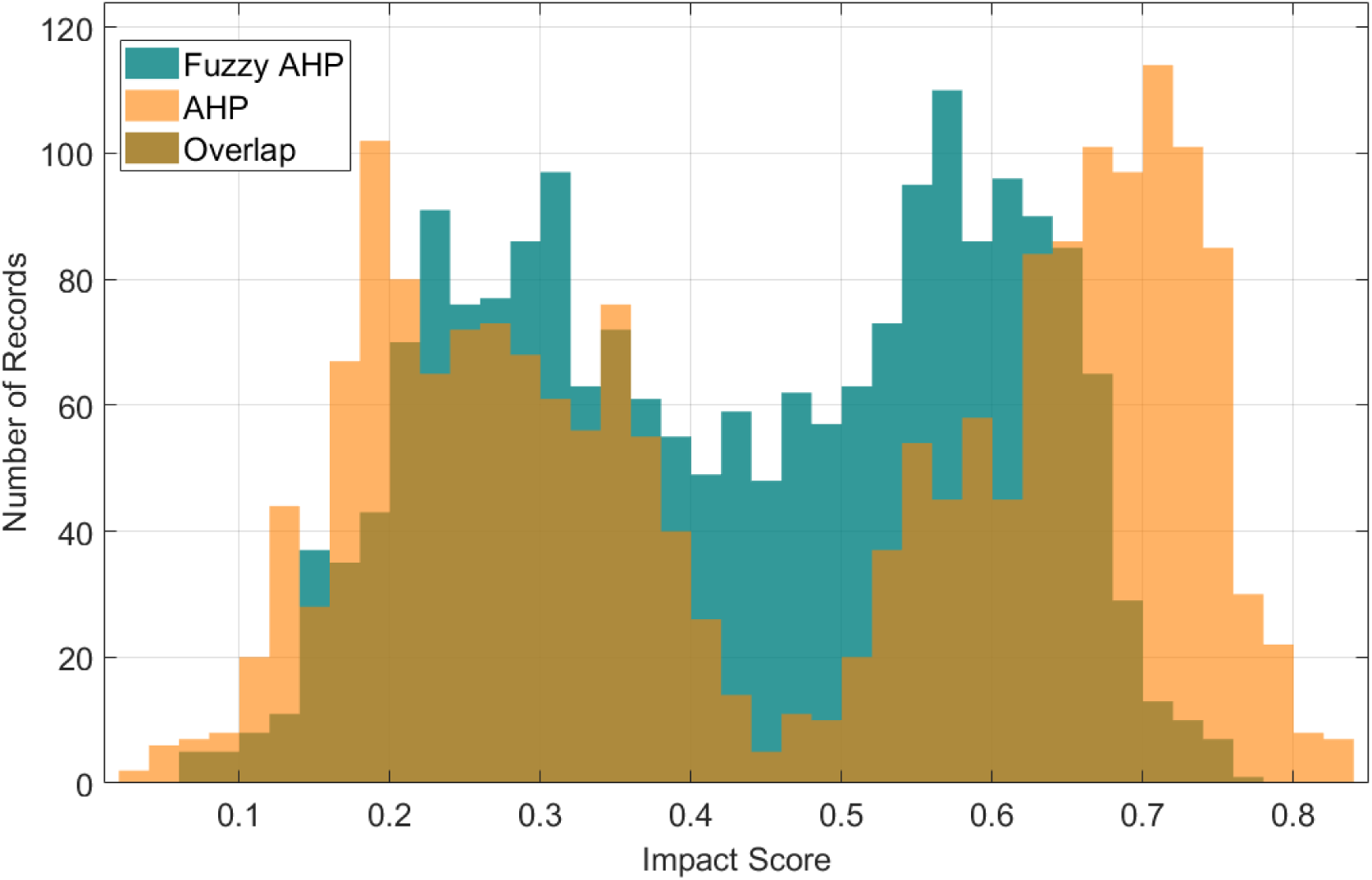
Distribution of overall impact index scores for agricultural incidents under AHP and Geometric-FAHP framework.

On the other hand, the AHP distribution exhibits a wider and more polarized structure, with a prominent peak in the 0.70 to 0.72 range. AHP assigns a larger number of incidents to both the lower (below 0.12) and upper (above 0.76) tails, indicating higher variance and stronger separation between low- and high-impact cases. While both methods show substantial overlap in the intermediate range (0.30 to 0.50), divergence is most apparent at the extremes. Essentially, the AHP approach functions as a contrast-enhancing mechanism that accentuates impact differentiation, whereas Geometric-FAHP acts as a smoothing mechanism that emphasizes gradual vulnerability transitions.

### 3.4. Geospatial Impact Density and Risk Visualization

The combined effects of incident frequency, outcome severity, and index-based weighting are most evident when examined spatially. While the preceding analyses focused on case-level behavior and mechanism-specific patterns, evaluating how the two weighting formulations perform at a broader regional scale is necessary before interpreting the continuous risk surfaces. To provide this, state-level summary statistics were compiled for both the AHP and Geometric-FAHP impact scores. This high-level comparison serves two purposes: to verify whether the structural differences between the weighting schemes persist after geographic aggregation and to establish the expected contrast and dispersion in the subsequent kernel density maps. Table 6 functions as a diagnostic step linking the index formulation to its spatial expression across the Midwest region.

**Table 6.**
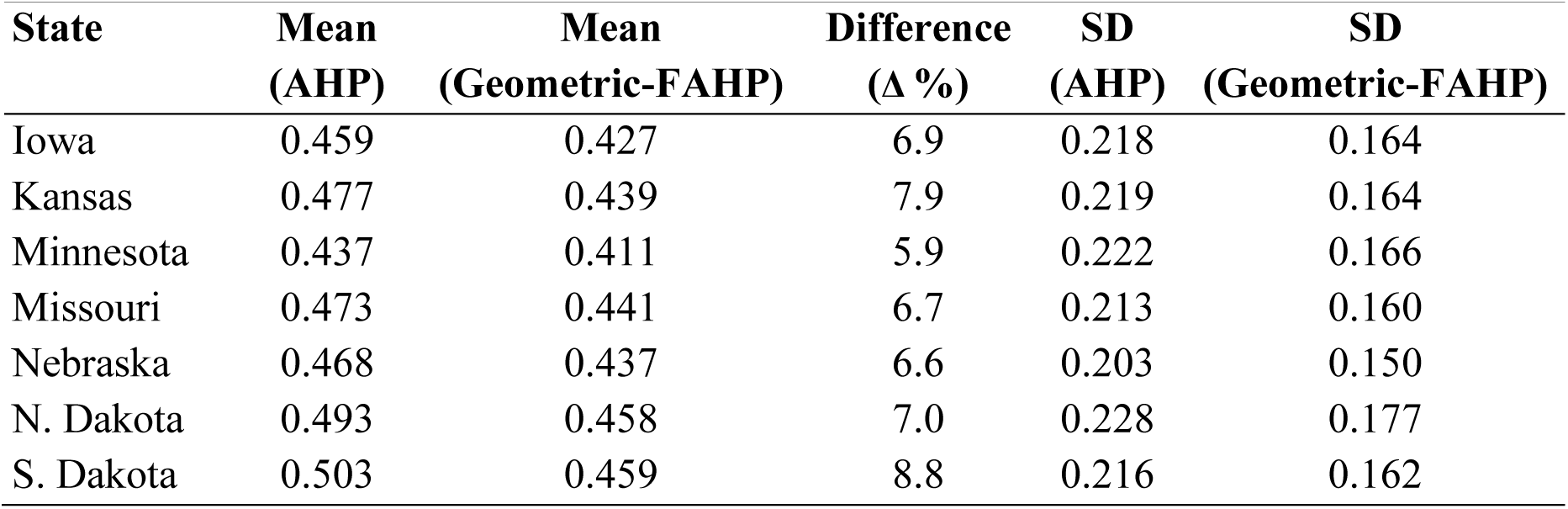
State-level summary statistics of AHP and Geometric-FAHP impact scores across the Midwest study region (SD: standard deviation)

| State | Mean<br>(AHP) | Mean<br>(Geometric-FAHP) | Difference<br>($\Delta$ %) | SD<br>(AHP) | SD<br>(Geometric-FAHP) |
| --- | --- | --- | --- | --- | --- |
| Iowa | 0.459 | 0.427 | 6.9 | 0.218 | 0.164 |
| Kansas | 0.477 | 0.439 | 7.9 | 0.219 | 0.164 |
| Minnesota | 0.437 | 0.411 | 5.9 | 0.222 | 0.166 |
| Missouri | 0.473 | 0.441 | 6.7 | 0.213 | 0.160 |
| Nebraska | 0.468 | 0.437 | 6.6 | 0.203 | 0.150 |
| N. Dakota | 0.493 | 0.458 | 7.0 | 0.228 | 0.177 |
| S. Dakota | 0.503 | 0.459 | 8.8 | 0.216 | 0.162 |

As shown in Table 6, the AHP formulation yields consistently higher mean impact scores than the Geometric-FAHP approach across all states, indicating that the crisp weighting structure retains stronger influence from severity-driven components. In contrast, the fuzzy formulation distributes weight more broadly across contextual and accessibility factors and therefore produces uniformly lower averages. This systematic reduction is reflected in the percent difference metric (Δ %), which quantifies the consistent gap (5.9–8.8%) between the AHP and Geometric-FAHP scores across states. The dispersion metrics follow the same pattern, with Geometric-FAHP exhibiting a lower standard deviation (SD) in all states. This reflects the compression of the score range and smoother regional differentiation, while AHP maintains greater variability consistent with its stronger sensitivity to high-consequence incidents. Although the relative ordering of states is broadly consistent under both methods, higher average impact levels are more pronounced in parts of the northern plains, while Minnesota occupies the lower end of the range and the central Midwest states fall in between. Given differences in incident volume among states, these patterns are interpreted as indicative rather than strictly rank-based, and they collectively highlight the contrasting behavior of the two weighting frameworks.

To examine the spatial behavior of the impact results, kernel density estimation is applied to generate continuous surfaces (Figure 6). Because the kernel outputs are based on normalized values, the color gradients represent relative clustering intensity rather than absolute incident counts. The maps should therefore be interpreted comparatively to identify where risk concentrates and how the weighting schemes modify the spatial pattern.

**Figure 6.**
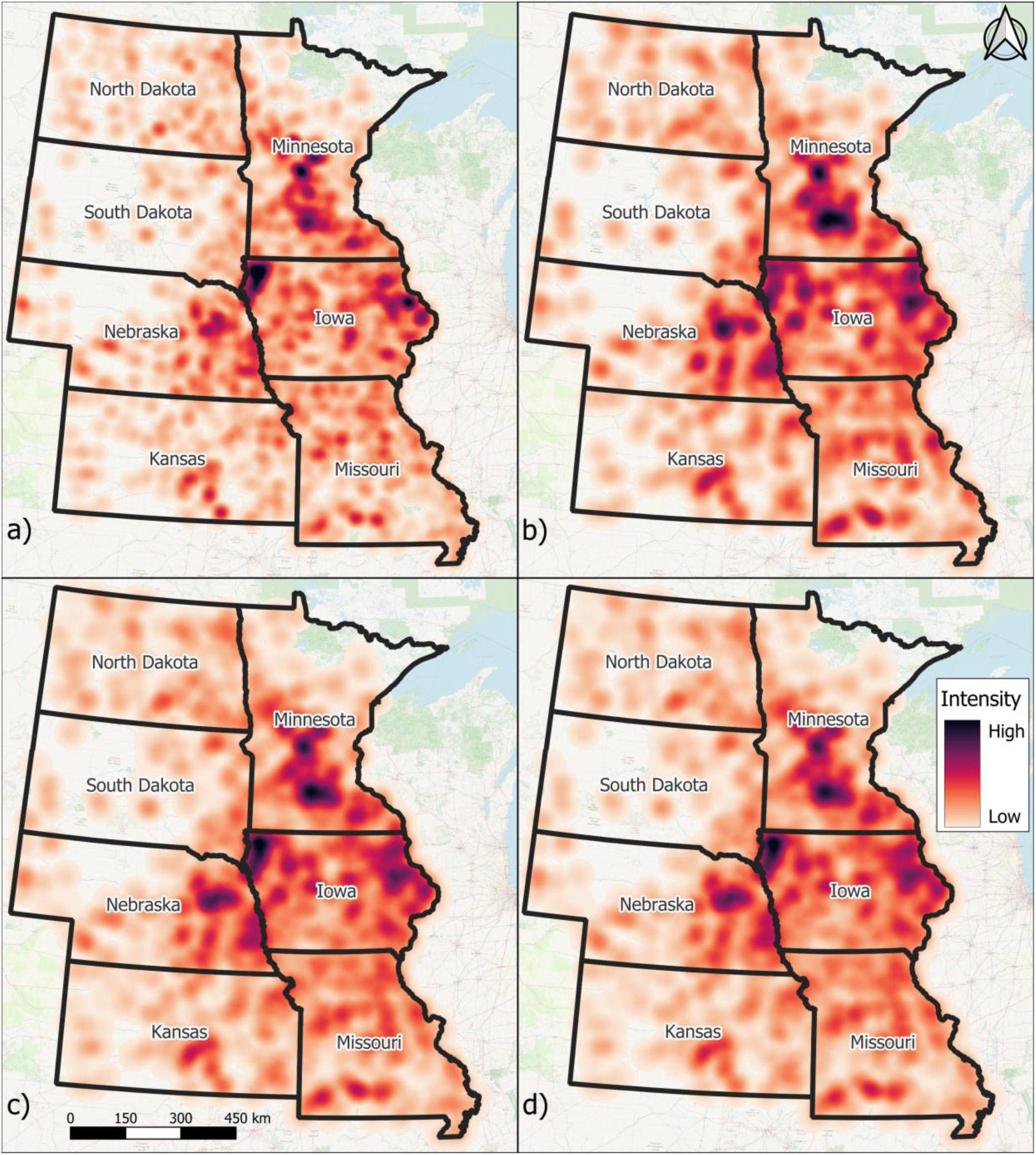
Quartic kernel density maps illustrating spatial patterns of agricultural incidents in the Midwest: a) unweighted incident density; b) fatality density; c) AHP-weighted impact density; and d) Geometric-FAHP–weighted impact density.

As shown in Figure 6a, the unweighted incident density surface represents the baseline exposure structure of agricultural activity across the Midwest. The most prominent feature is the broad and continuous core in northwestern Iowa, which forms the strongest concentration of incident frequency in the region. Additional activity appears as localized spikes in central Minnesota and in eastern Iowa near and north of the Dubuque area, while eastern Nebraska shows moderate clustering near the Iowa border. Southern Missouri and southern Kansas contain scattered mid-level spikes, and both Dakotas remain comparatively diffuse. Overall, the pattern closely follows the geographic footprint of agricultural operations, indicating that the unweighted surface primarily reflects where incidents occur most frequently.

In Figure 6b, the fatality density pattern shows a wider regional spread than the total incident map. Elevated intensity extends across central and northern Iowa, while the central–southern portion of Minnesota becomes more prominent relative to the total incident distribution. Eastern Nebraska shows increased clustering near the Iowa border, and southern Missouri develops additional pockets of elevated intensity. South Kansas and eastern South Dakota show minor increases but remain below the primary high-intensity zones, and much of western South Dakota continues to display weak patterns. This shift from a single dominant exposure core toward a broader distribution indicates that fatal outcomes are not strictly proportional to incident frequency, consistent with the earlier statistical results.

Figure 6c shows that the AHP-weighted impact distribution generally follows the fatality pattern but introduces stronger spatial consolidation. Northwestern Iowa remains the dominant hotspot and appears both broader and more intensely concentrated than in the base and fatality maps. Across Iowa, the impacted area becomes more spatially continuous, particularly in the northeast, while central and southern Minnesota maintain strong influence and the southeastern Minnesota–Iowa corridor becomes more pronounced. In eastern Nebraska, previously separated clusters consolidate into two denser cores along the Iowa border and the central-eastern region. Kansas largely preserves its fatality-based pattern but shows stronger mid-level clustering near the Missouri boundary, and Missouri becomes more spatially even through its central and southern portions. North and South Dakota remain comparatively light with diffuse patterns. These behaviors indicate that the AHP formulation accentuates locations where elevated exposure and high-severity mechanisms coincide, producing sharper spatial differentiation than the fatality surface alone.

Finally, in Figure 6d, the Geometric-FAHP surface retains the same overall hotspot hierarchy observed under AHP but with lower contrast and smoother spatial transitions. Northwestern Iowa remains the dominant hotspot and becomes more clearly distinguished relative to eastern Iowa, while most other areas appear lighter and more spatially diffuse. This behavior is consistent with the earlier statistical evidence showing lower variance under the fuzzy formulation. Rather than introducing new hotspot locations, the fuzzy weighting reduces intensity gradients and produces a more continuous regional pattern of impact. The similarity in spatial ordering between the two weighted surfaces supports that the primary risk structure is stable, while the contrast differences reflect the distinct behavior of the weighting schemes.

Overall, the sequence of surfaces shows consistent progression from exposure-based patterns to severity- and index-informed impact representation. The unweighted map identifies where incidents are most frequent, the fatality surface introduces outcome-based differentiation, and the AHP and Geometric-FAHP formulations further integrate multi-criteria effects. Within this progression, AHP produces stronger spatial contrast, whereas the fuzzy approach yields a smoother regional pattern, providing complementary perspectives on agricultural risk across the Midwest.

## 4. Conclusion

This study conducted a multi-year geospatial analysis and multi-criteria risk assessment of agricultural incidents across seven U.S. Midwest states from 2012 to 2023. By integrating longitudinal incident surveillance data with environmental and infrastructural layers, the research established that incident frequency and outcome severity do not share identical spatial footprints across the region. While overall incident density is heavily concentrated in the intensive row-crop cores of northwestern Iowa and central Minnesota, fatal outcomes exhibit a broader and more fragmented distribution. This spatial divergence confirms that relying solely on raw incident counts would significantly underrepresent localized high-consequence areas, as total incident frequency primarily reflects baseline agricultural exposure rather than inherent danger.

The analysis identified a clear separation between frequent but survivable events and high-consequence mechanisms that govern loss-of-life outcomes. Roadway and entanglement incidents occur more routinely but are characterized by a lower lethality range. Conversely, equipment-related categories remain consistently above the regional average fatality rate of 54.1%, with skid-loader incidents emerging as the most lethal mechanism at 76.9%. High fatality proportions are also observed for tractors (58.6%), grain bins (60.0%), and ATV/UTV events (61.2%), which often involve heavy machinery, rollovers, or confined spaces. Demographic patterns further clarify this risk distribution, showing that hazardous agricultural activities remain heavily concentrated among male workers, who represent 85% of fatal outcomes. Furthermore, the fatality rate increases with advancing age, with the 60 to 70 age group bearing the highest fatal burden in the dataset.

Methodologically, the comparative application of AHP and Geometric-FAHP provided complementary perspectives on regional agricultural risk. The AHP approach functions as a contrast-enhancing mechanism, highlighting impact differentiation and emphasizing localized, high-impact hotspots where catastrophic incidents dominate. In contrast, the Geometric-FAHP framework acts as a smoothing mechanism that emphasizes gradual vulnerability transitions and identifies broader regional risk structures that might otherwise be obscured by severity-driven extremes. By explicitly linking incident locations with survivability factors, such as proximity to emergency medical services and cellular infrastructure, this research provides an evidence base for targeting safety interventions and addressing the digital divide in rural emergency response planning.

Despite the robustness of the multi-state database, several limitations persist, including the fragmented nature of current injury surveillance and the likelihood of underreporting when relying on single data sources. Furthermore, approximately 4% of the total records lacked sufficient location precision for high-resolution spatial modeling. Future research should focus on integrating more granular human factor data and exploring the temporal evolution of these risk surfaces in relation to evolving agricultural practices and climate variability. Ultimately, this research moves beyond simple frequency metrics to offer a novel, quantitative mechanism to assess the multidimensional impact and severity of agricultural incidents across the Midwest region.

## Author Contributions

E.D.: conceptualization, methodology, validation, data curation, visualization, formal analysis, and writing—original draft preparation; O.M.: conceptualization, methodology, software, data curation, and visualization; I.D.: writing—review and editing, validation, supervision, project administration, and funding acquisition. All authors have read and agreed to the published version of the manuscript.

## Funding

This research received no external funding.

## Data Availability Statement

The data used in this study are available in the Central States Center for Agricultural Safety and Health (CS-CASH) under the Enhanced Farm and Agricultural Injury Media Monitoring Program at https://www.unmc.edu/publichealth/cscash/resources/fatalities-injuries-by-state.html (accessed on March 12, 2025).

## Acknowledgment

The authors thank Enes Esref Onder for his valuable contributions.

## Conflicts of Interest

The authors declare no conflicts of interest.

This manuscript is an medRxiv preprint and has been submitted for possible publication in a peer reviewed journal. Please note that this has not been peer-reviewed before and is currently undergoing peer review for the first time. Subsequent versions of this manuscript may have slightly different content.

## Notes

### Competing Interest Statement

The authors have declared no competing interest.

